# Decomposing Participatory Surveillance Symptom Time Series to Track Respiratory Infections: A Cross-Country Evaluation Using Non-Negative Matrix Factorization

**DOI:** 10.64898/2026.03.30.26349719

**Authors:** Gesa Carstens, Mattia Mazzoli, Nicolo Gozzi, Albert Jan van Hoek, Daniela Paolotti

## Abstract

**Background:** The annual respiratory season in Europe is marked by the co-circulation of multiple respiratory pathogens, such as influenza viruses, rhinoviruses, and coronaviruses. Effective surveillance is necessary but hampered by heterogeneity of case definitions and limited pathogen specificity in existing systems. This study aims to detect pathogen-specific signals in the participatory surveillance of the Netherlands using a sub-set of samples with virological detection. Additionally, we explore a method to use the findings in the Netherlands to enhance the virological interpretation of participatory surveillance data in Italy.

**Methods:** We analyzed symptom data collected through a participatory surveillance platform in the Netherlands and Italy over five years (2020–2025). Symptom-by-week matrices from the Dutch cohort were aggregated into syndromes and their associated time series using Non-negative Matrix Factorization (NMF). We compared the respective time series of the syndromes with influenza virus, SARS-CoV-2, seasonal coronaviruses, RSV, and rhinovirus incidence estimated from nose- and throat swabs of a subsample of symptomatic participants of the participatory surveillance platform in the Netherlands. We tested the transferability of these components by applying the Dutch-derived components to describe Italian symptom data and extract respective incidences.

**Results:** NMF identified eight symptom clusters in the Dutch cohort, one aligning with SARS-CoV-2, one aligning with rhinovirus and a third component aligning with influenza virus, RSV and seasonal incidences estimated from collected nose- and throat swabs. Transferring Dutch-derived symptom clusters to Italian data showed consistency in key components between Dutch and Italian cohorts, particularly those associated with SARS-CoV-2.

**Conclusion:** This study demonstrates that unsupervised symptom decomposition can identify co-circulating respiratory pathogens trends from syndromic surveillance data, providing timely pathogen circulation insights.

**Graphical abstract:** 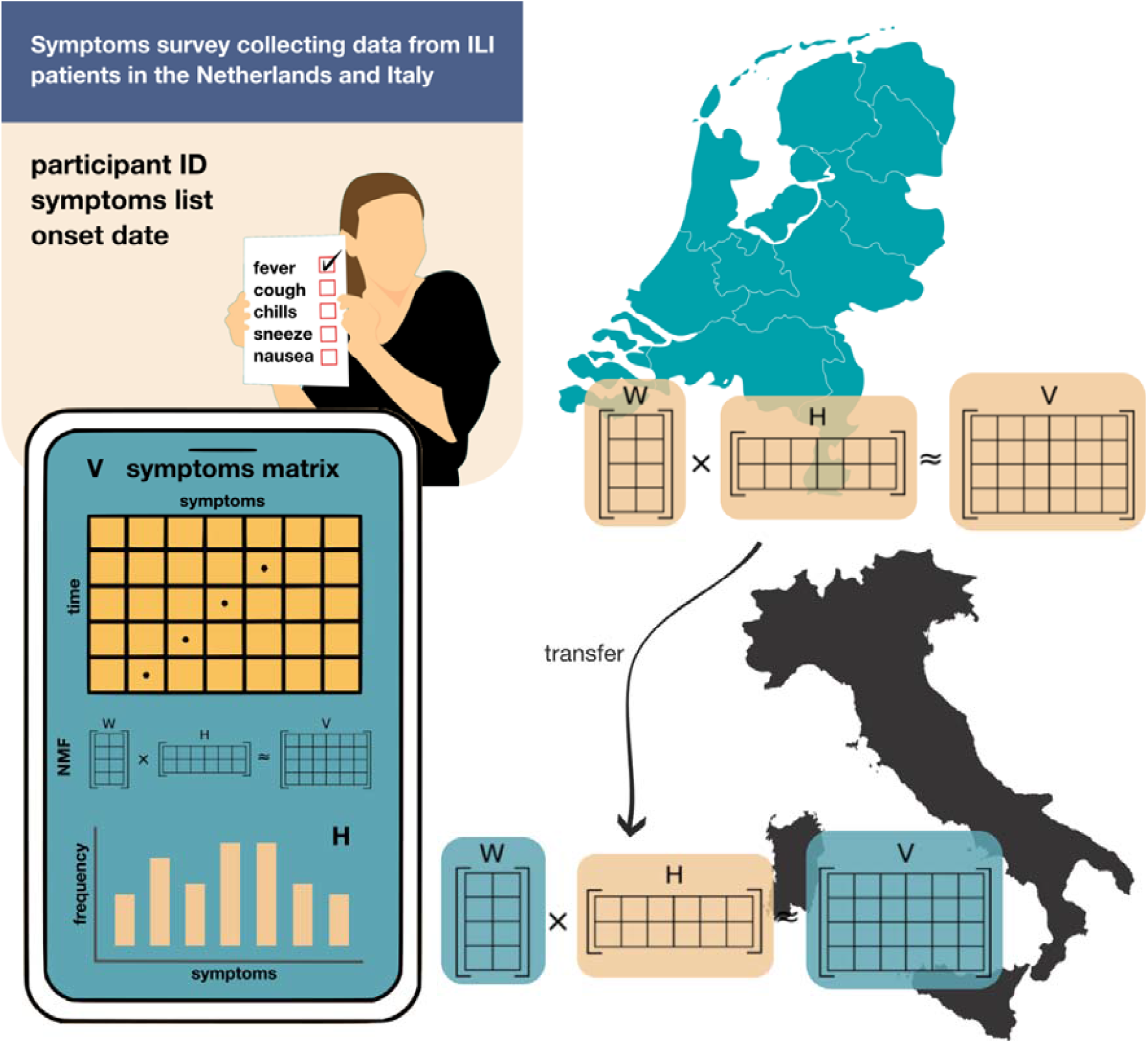

## INTRODUCTION

The yearly respiratory season in Europe is marked by the co-circulation of a variety of respiratory pathogens, including influenza viruses, rhinoviruses, adenoviruses, and coronaviruses. These pathogens cause a range of general and respiratory symptoms, with illness severity spanning from mild cases that resolve within a few days to severe disease that may necessitate hospitalization or can even lead to death, particularly among children, the elderly, and other at-risk groups (1). Effective surveillance of these infections is essential for public health action, yet current systems across Europe are heterogeneous and face important limitations.

Traditional virological surveillance, typically conducted via sentinel surveillance systems where GPs and hospitals collect samples from symptomatic patients, offers high specificity but is resource-intensive and limited in coverage (2). However, most countries rely on syndromic surveillance systems – such as tracking influenza-like illness (ILI) consultation rates among GPs or in the hospital – that provide broader coverage and timely insights yet lack pathogen-specific detail and standardized case definitions. Both surveillance systems have proven valuable during seasonal epidemics and public health emergencies, like the 2009 H1N1 pandemic (3), but their respective limitations highlight the importance of integrating both syndromic and virological data to achieve comprehensive understanding of respiratory disease dynamics across Europe.

To complement these traditional systems, scalable participatory surveillance platforms such as Influenzanet in Europe have emerged as transnational efforts (4,5). Influenzanet encompasses multiple platforms in European countries inviting individuals to voluntarily report their symptoms via weekly online questionnaires, allowing for timely and potentially more representative data collection overcoming healthcare-seeking behavior biases of traditional systems (6). Previously, incidence trends derived from Influenzanet data have been shown to align well with traditional sentinel surveillance (2,5). The harmonized structure of these platforms across Europe creates unique opportunities for cross-country collaboration and comparative analyses recognizing the cross-border transmission of respiratory pathogens. Recent innovations, such as the introduction of a self-swabbing component to the Dutch Infectieradar platform in 2022, now allows to integrate laboratory-confirmed virological data with syndromic surveillance at the individual level.

Our previous work introduced an unsupervised framework to derive an ILI signal from weekly symptom reports, enabling robust tracking of ILI incidence across countries and seasons (7). This approach assumes that symptoms co-vary over time in ways that reflect the underlying circulation of respiratory syndromes. With the recent availability of virological data within the Dutch participatory surveillance platform, we can extend this framework to move beyond an ILI signal and instead identify pathogen-specific signals represented by latent symptom components. We additionally explored whether components trained on data from one country, where pathogen circulation within the online participatory platform is known, can be exported to another country without self-sampling of the participants. If transferable, these components would allow symptom-based surveillance systems elsewhere to approximate pathogen-specific trends using laboratory data from a single country as the calibration source.

## METHODS

### Data collection

#### Participatory surveillance data

For this study we used data from the Italian Influweb online platform, which has been operational since winter 2008, and the Dutch Infectieradar platform, which was launched more recently, in November 2020. Details of the design and methodology of Infectieradar and Influweb can be found in (4,8). Briefly, voluntary participants complete a background questionnaire upon registration, reporting their sociodemographic details and medical history. Participants are then asked weekly to fill out a brief questionnaire to report whether they had experienced any of 22 symptoms (fever, chills, rhinitis, sneeze, sore throat, cough, dyspnea, headache, pain, chest pain, asthenia, anorexia, sputum, watery eye, nausea, vomiting, diarrhea, abdominal pain, loss of smell, loss of taste, nosebleed, sudden onset (i.e., symptom onset within 24 hours)). Those who report symptoms are prompted to answer additional questions about the onset date of their symptoms, the severity of the illness, healthcare-seeking behavior, testing and medication use. If participants report symptoms in consecutive weeks, they are asked whether the current symptoms were part of the same infection episode as the previous ones, allowing for data collection on the same infection episode over multiple weeks.

For our analyses we included participants from the participatory surveillance platforms who filled in a total of at least two weekly questionnaires during their participation to exclude participants who registered to the study but only participated once or never. If a participant submitted more than one weekly questionnaire in a given week, we included only the most recent one. To ensure the reports reflect the complete spectrum of symptoms during the acute phase of infection, we only included symptoms reported within 3-14 days of symptom onset.

#### Italian sentinel virological surveillance

The fraction of positive tests of respiratory viruses in Italy was estimated through virological confirmation conducted among patients presenting to their GP with ILI or from hospital-based surveillance (9). National-level virological prevalence data among ILI patients in Italy have been available for influenza virus A and B since October 2019. Since October 2021, data have also been available for adenovirus, bocavirus, influenza virus A and B, seasonal coronavirus, human metapneumovirus, parainfluenza virus, respiratory syncytial virus, rhinovirus, and SARS-CoV-2.

### Laboratory assays of Infectieradar self-swabs

From October 2022 the Dutch participatory surveillance platform Infectieradar was extended by the addition of a self-testing component (8). Participants taking part in this study receive SARS-CoV-2 self-tests along with materials for a nose and throat sample (NTS). Upon reporting ARI symptoms (i.e., reporting at least one of the following symptoms within 5 days of symptom onset: a sore throat, rhinorrhea, a runny nose, or dyspnea) participants could be invited to send a NTS to our laboratory. Upon arrival, RNA is extracted from the NTS using MagNApure 96 (MP96) (Roche) and extracted RNA is subsequently analyzed using SARS-CoV-2 specific RT-PCR as described (10) and multiplex real-time PCR (RespiFinder 2SMART, PathoFinder, the Netherlands). Samples are tested for the presence of the following respiratory viruses: SARS-CoV-2, influenza virus A, influenza virus B, RSV-A, RSV-B, human metapneumovirus, rhino-/enterovirus, adenovirus, parainfluenza-1, parainfluenza-2, parainfluenza-3, parainfluenza-4, bocavirus, seasonal human coronavirus NL63, seasonal human coronavirus HKU1, seasonal human coronavirus OC43, seasonal human coronavirus 229E and MERS-CoV. Although the multiplex real-time PCR technology used is not officially able to distinguish between rhinovirus and enterovirus, further in-house typing of a representative subset of samples indicated that most of these samples were rhinovirus positive. Therefore, we refer to them as rhinovirus in the main text and results.

#### Weekly pathogen incidence

The weekly incidence of each pathogen in the Netherlands was estimated by multiplying the proportion of samples testing positive for that pathogen in a given week by the proportion of participants reporting acute respiratory infection symptoms (as at least one of the following: cough, sore throat, dyspnea, or runny nose) during the same week. Weekly SARS-CoV-2 incidence was estimated separately, by dividing the number of positive PCR or self-test reports by the number of active participants for each week. The weekly incidence for rhinovirus, influenza virus, seasonal coronaviruses and RSV was available for October 2022 to May 2025, while the weekly incidence for SARS-CoV-2 was available from November 2020 to May 2025. Weekly incidence of each pathogen in Italy was estimated by multiplying the proportion of positive tests in a given week by the ILI incidence reported by national sentinel surveillance.

### Statistical analysis

#### Input data

Similarly to previous work (7), we hypothesize that among all reported symptoms, certain symptom groups represent the symptomatic expression of underlying infections experienced by participants of the participatory surveillance platforms.

We build two separate input matrices using the Dutch Infectieradar data and the Italian Influweb data from November 2020 to May 2025. To ensure the reports reflect the complete spectrum of symptoms during the acute phase of infection, we only included symptoms reported within 14 days of symptom onset. Each symptom was recorded as either present or absent, and weekly symptom counts were aggregated across participants. To account for participation variability, we calculated symptom incidences within each study population by normalizing the counts based on the number of active participants each week. Participants were defined as active if they submitted at least one weekly questionnaire in the two weeks prior and two weeks after the current submission. To ensure that more commonly reported symptoms do not skew the results we normalized the symptoms across weeks so that each symptom has a weekly incidence between 0 and 1. The final incidence matrix is represented as:

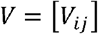

Where *V*_*ij*_ denotes the incidence of symptom *j* during week *i*. This matrix can be linearly decomposed using NMF to extract hidden symptom groups representing the infections caused by circulating respiratory pathogens.

#### Non-negative matrix factorization

NMF is an unsupervised method for extracting meaningful patterns from data by decomposing it into a set of nonnegative features (i.e., components) and their associated time series (11). The nonnegativity constraint ensures that all components remain interpretable, as negative values would be meaningless in this context. This property makes NMF particularly well-suited for applications where data naturally consists of additive parts, such as symptom prevalence in disease surveillance.

NMF seeks to approximate a given nonnegative matrix V by factorizing it into the product of two lower-dimensional nonnegative matrices:

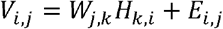

Where:

- *V* is the observed data matrix with *i* rows (i.e., weeks) and *j* columns (i.e., symptoms);
- *W* contains the syndromic spectrum with *j* rows and *k* columns, where each column represents a latent component and rows encode the symptoms in each component;
- *H* is the coefficient matrix with *k* rows and *i* columns, describing how strongly each component contributes to the observed data at week *i*;
- *E* is the approximation error with *i* rows and *j* columns;
- *k* is the number of latent components.

#### Matrix decomposition by NMF

The decomposition of the input matrix is achieved by solving the following optimization problem:

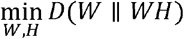

Where *D* (*W* ║ *WH*) is a divergence function measuring the difference between *V* and its approximation *WH*. We used the Kullback-Leibler (KL) divergence loss function (12) defined as:

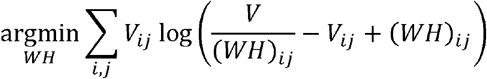

To improve convergence and interpretability, we initialize *W* and *H* using Non-negative Double Singular Value Decomposition (NNDSVD) (13). This method provides a structured initialization by approximating principal components while enforcing non-negativity.

Through this decomposition, each column of *W* represents a distinct syndrome spectrum, while each row of *H* quantifies how strongly each component contributes to the overall signal per week. It is important to note that the symptom distribution within each component does not directly reflect the absolute occurrence of symptoms in individuals infected with a specific pathogen. Instead, it highlights differences between components and should be interpreted in relation to the other extracted components. Since NMF seeks to decompose the observed data into additive parts, symptoms are distributed across components in a way that best explains the overall variance. Therefore, the symptom clusters of each component reflect pathogen specific average syndromic spectra in the population, and in this sense should not be interpreted as probabilistic case definitions.

#### Selection of the optimal number of components

An important consideration in NMF is determining the optimal number of components for the decomposition. In our case, the incidence matrix *V* consists of *j* = 22 symptoms, establishing 22 as the upper bound for the number of components. The goal is to select a number of components that effectively capture the underlying structure of the original incidence matrix while avoiding overfitting. To achieve this, we evaluated the decomposed matrix for each *k*- component solution and assessed model fit using the corrected Akaike Information Criterion (AICc). The AICc was chosen to account for a small sample size, ensuring a balance between model complexity and predictive accuracy.

#### Component extraction from the Dutch data

First, we used the preprocessed input matrix containing the Dutch Infectieradar data to determine the optimal number of components as described above. Next, we applied NMF to the Dutch input data to extract the latent components from the symptom matrix. From the extracted components, we identified those most strongly correlated with weekly pathogen incidence through Pearson correlation analysis. Specifically, for SARS-CoV-2, influenza virus, and rhinovirus, RSV, and seasonal coronaviruses we identified the components with the highest Pearson correlation coefficient (>0.65) with the weekly incidence of these specific pathogens as measured by the submitted NTS from symptomatic participants each week.

#### Transferring components across countries

Next, we informed the NMF using all components detected from the Dutch cohort, to infer weekly pathogen incidence from the Italian symptom reports using the Italian input matrix. For this, the symptoms-by-week matrix from the Italian Influweb cohort and the *W* matrix of components’ symptomatic spectra extracted from the Dutch cohort were used to infer the *H* matrix encoding the components’ time series, i.e. components incidence, at weekly scale:

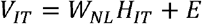

Where:

- *V*_*IT*_ is the observed Italian symptom matrix
- *W*_*NL*_ contains the syndromic spectrum extracted from the Dutch NMF
- *H*_*IT*_ is the coefficient matrix, describing the component incidence
- *E* is the approximation error

Next, we calculated the Pearson correlation of the inferred *H*_*IT*_ matrix and the pathogen time series reported by the Italian national virological surveillance (9).

## RESULTS

### Study Population

From November 2020 to May 2025, a total of 39,014 participants were enrolled in the Dutch Infectieradar system, contributing 2,803,722 weekly symptom reports. Likewise, in the Italian Influweb system 5873 participants reported 30,886 weekly symptom reports since November 2011 to May 2025. Sociodemographic characteristics of the study population are summarized in **Table 1**. The Dutch and Italian cohorts were overrepresented by females, a trend that was even more pronounced among those submitting NTS in Infectieradar. Additionally, the majority of participants were aged 50 years or older and reported having a high educational level.

**Table 1.** Participant characteristics of Infectieradar and Influweb.

|  | Infectieradar<br>N = 39,014 | Influweb<br>N = 5,873 |
| --- | --- | --- |
| **Sex** |  |  |
| female | 22,960 (59%) | 2,532 (43.1%) |
| male | 15,830 (41%) | 3,084 (52.5%) |
| Unknown | 224 (0.6%) | 257 (4.4%) |
| **Age group (years)** |  |  |
| <25 | 1,968 (5.1%) | 944 (16.1%) |
| 25-39 | 7,211 (19%) | 1,547 (26.3%) |
| 40-49 | 6,915 (18%) | 1,141 (19.4%) |
| 50-64 | 14,371 (37%) | 1,149 (19.6%) |
| 65+ | 8,404 (22%) | 425 (7.2%) |
| Unknown | 145 (0.4%) | 667 (11.4%) |
| **Education level** |  |  |
| lower/none | 912 (2.4%) | 2,459 (41.9%) |
| middle | 14,613 (38%) | 1,657 (28.2%) |
| higher | 22,773 (59%) | 1,757 (29.9%) |
| Unknown | 716 (1.8%) |  |
| **Smoking** | 4,222 (11%) | 1,059 (18.0) |
| **Comorbidities** |  |  |
| Allergy | 14,486 (37%) | 1,510 (25.7%) |
| Asthma | 3,020 (7.8%) | 207 (3.5%) |
| Diabetes | 1,457 (3.7%) | 119 (2.0%) |
| Chronic lung disease | 986 (2.5%) | 55 (0.9%) |
| Cardiovascular disease | 3,4394 (8.7%) | 484 (8.2%) |

### Component extraction from Dutch data

To determine the optimal number of components k, we computed the AICc score for a range of NMF components applied to the incidence matrix of Infectieradar. The AICc score balances an increasing penalty for higher number of k-components and their accuracy at reproducing the observed symptoms trends (see Methods). The AICc was minimized at, suggesting that eight latent components effectively captured the underlying structure of the dataset (**Supplemental Figure 1**). The eight extracted components represent distinct syndromic spectra present in the dataset. We represent their associated symptom loadings and time trends in **Figure 1**.

**Figure 1:**
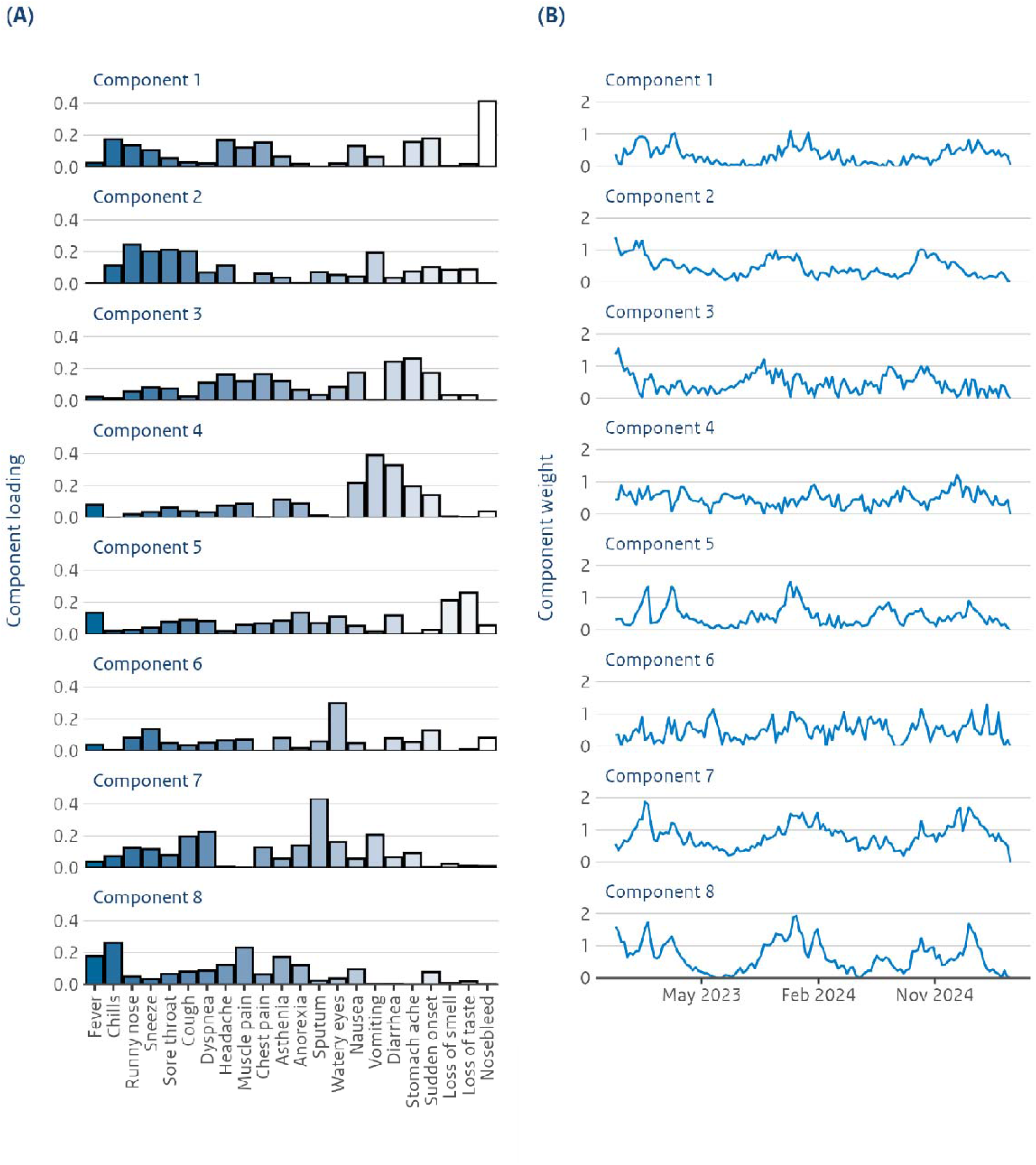
Extracted components from the Dutch Infectieradar data and their time-series. (A) Syndromic spectrum of the components (W matrix). The bars are colored for readability purposes only. (B) The respective time series of each component (H matrix).

Using Pearson correlation analysis, we assessed the association between the temporal dynamics of each component and weekly pathogen incidence rates derived from laboratory-confirmed cases of the self-testing study and SARS-CoV-2 positive self-test reports (**Table 2**).

**Table 2.** Pearson correlation of the NMF components and the weekly pathogen incidence from the Netherlands.

| Component | Influenza virus | SARS-CoV-2 | Rhinovirus | seasonal<br>coronaviruses | RSV |
| --- | --- | --- | --- | --- | --- |
| 1 | 0.58<br>(p = <0.001) | 0.43<br>(p = <0.001) | 0.3<br>(p = <0.001) | 0.6<br>(p = <0.001) | 0.66<br>(p = <0.001) |
| 2 | -0.1<br>(p = 0.26) | 0.45<br>(p = <0.001) | 0.88*<br>(p = <0.001) | -0.08<br>(p = 0.34) | 0.38<br>(p = <0.001) |
| 3 | -0.24<br>(p = <0.01) | 0.28<br>(p = <0.001) | 0.43<br>(p = <0.001) | -0.38<br>(p = <0.001) | -0.11<br>(p = 0.22) |
| 4 | 0.38<br>(p = <0.001) | -0.22<br>(p = <0.001) | -0.13<br>(p = 0.14) | 0.38<br>(p = <0.001) | 0.27<br>(p = <0.01) |
| 5 | 0.39<br>(p = <0.001) | 0.76*<br>(p = <0.001) | 0.36<br>(p = <0.001) | 0.27<br>(p = <0.01) | 0.52<br>(p = <0.001) |
| 6 | 0.09<br>(p = 0.32) | 0.17<br>(p = 0.011) | 0.2<br>(p = 0.021) | 0.19<br>(p = 0.029) | 0.12<br>(p = 0.17) |
| 7 | 0.68*<br>(p = <0.001) | 0.27<br>(p = <0.001) | 0.28<br>(p = <0.01) | 0.73*<br>(p = <0.001) | 0.7*<br>(p = <0.001) |
| 8 | 0.48<br>(p = <0.001) | 0.7<br>(p = <0.001) | 0.55<br>(p = <0.001) | 0.34<br>(p = <0.001) | 0.51<br>(p = <0.001) |

**Note:** \* indicate components with the highest correlation with estimated weekly pathogen incidence within Infectieradar participants

We found one component with a high correlation (*r* = 0.76, p < 0.001) with the weekly SARS-CoV-2 incidence, indicating that this component might capture the population-level symptomatic expression of participants infected with SARS-CoV-2 (**Figure 2**). The syndromic spectrum of this component was dominated by loss of smell and taste, fever, cough, anorexia and watery eye. The timing of the SARS-CoV-2 peaks among Infectieradar participants aligned well with the component time series, except in early 2025. During that time the percentage of SARS-CoV-2 reports was very low in the Infectieradar population, while the time series of the component shows a peak in February 2025.

**Figure 2.**
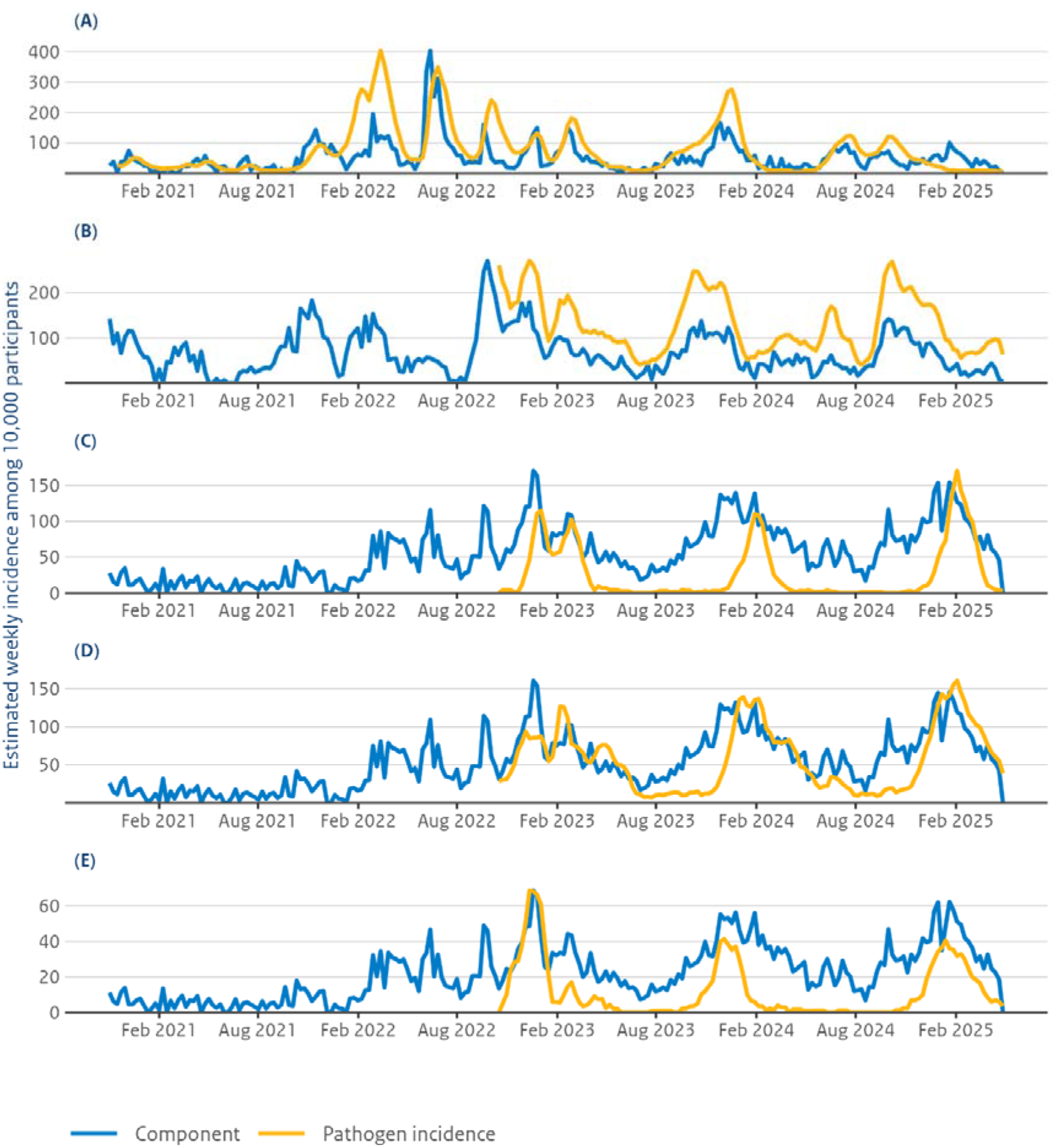
Time series of the pathogen-associated components in Infectieradar. (A) Comparison of the time series of the SARS-CoV-2 component and the weekly percentage of participants reporting a positive self-test or PCR test. (B) Comparison of the time series of the rhinovirus component and the weekly percentage of rhinovirus samples detected in submitted nose- and throat samples of Infectieradar participants multiplied by the percentage of new acute respiratory infections per week. Comparison of the time series of the component correlating with the weekly percentage of (C) influenza virus, (D) seasonal coronavirus and (E) RSV samples detected in submitted nose- and throat samples of Infectieradar participants multiplied by the percentage of new acute respiratory infections per week. For all figures the components magnitude was rescaled to match maximum and minimum of reported incidence. Time series of the pathogen incidence were smoothed a with a rolling average covering three weeks.

Next, another component demonstrated a strong correlation with laboratory-confirmed cases of rhinovirus (*r* = 0.88, p < 0.001). The temporal trends showed clear alignment, with the component peaks mirroring those of laboratory-confirmed rhinovirus incidence in Infectieradar participants, therefore we assume that this component reflects a rhinovirus-specific signal in the symptom data (**Figure 2**). The syndromic spectrum for this component emphasized a runny nose as a key feature, along with vomiting and several respiratory symptoms, such as sneezing, a sore throat, and cough. Interestingly, fever is absent in this component.

Lastly, we found a component not correlating with one specific pathogen but showing high correlations with influenza virus (*r* = 0.68, p < 0.001), seasonal coronaviruses (*r* = 0.73, p < 0.001), and RSV (*r* = 0.7, p < 0.001). The syndromic spectrum is dominated by sputum, dyspnea, and cough.

### Transferring components across countries

To test the generalizability and transferability of the method across countries, we applied the 8 components extracted from the Dutch cohort to inform the NMF method on the Italian symptoms matrix and inferred the incidence trends of the same pathogens among the Italian cohort (**Figure 3**). We find some Dutch components well correlating with incidence of SARS-CoV-2, rhinovirus, influenza virus and RSV from the Italian surveillance system (**Supplemental Table S1**). Not all year’s magnitudes match well with the reported incidence reflected also in the differences in correlation; this could be due to a substantial variation of participants in the Italian cohort that decreased after the end of the pandemic and the differences in the underlying source populations of the Influweb population and the national surveillance system. While the former contains mainly symptom reports from generally mild infections, the latter reflects respiratory virus circulation among ILI-patients at the GP or hospital. Virological validation of the components in the Italian cohort is therefore not possible.

**Figure 3:**
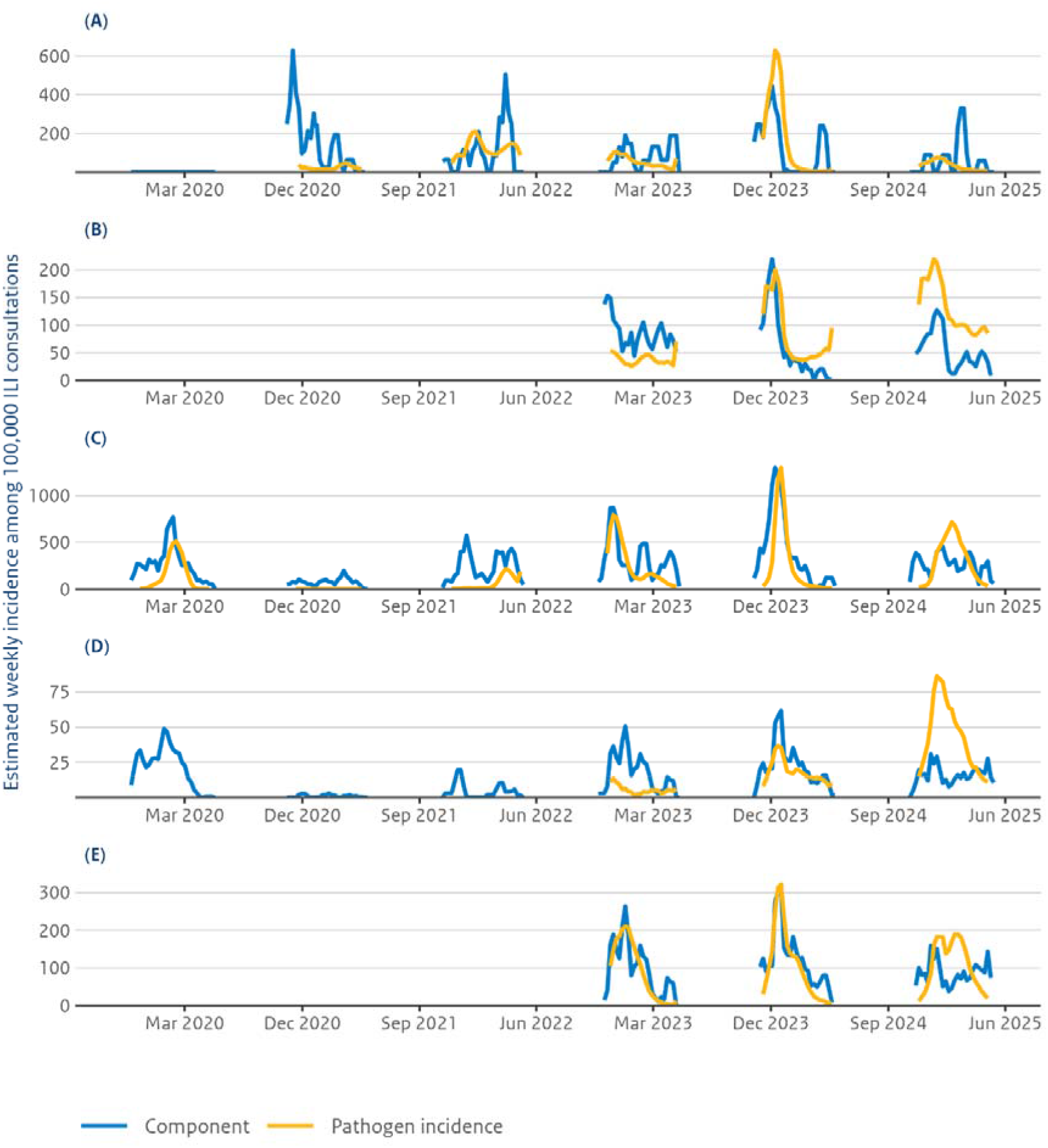
Time series of the Dutch components in Italy compared to national sentinel surveillance. Component time series compared to the national sentinel surveillance of (A) SARS-CoV-2, (B) rhinovirus, (C) influenza virus, (D) seasonal coronaviruses, and (E) RSV. Incidences for each virus were estimated as the product of ILI incidence modulated by the respective pathogen incidence among ILI patients measured from virological surveillance. Components magnitude was rescaled to match maximum and minimum of reported incidence. Time series of the pathogen incidence were smoothed a with a rolling average covering three weeks.

### Comparison of Dutch and Italian NMF components

We hypothesized a scenario in which we do not have access to NMF detected components from the Dutch case and we re-run NMF on the Italian symptoms’ matrix. The AICc indicated for this input matrix the optimal number of components to capture the variation in the data to be 6. We systematically compared the syndromic spectra of the Dutch and Italian components to check for consistency among detected syndromes (**Figure 4**). We observe a good consistency among some of the components detected in the two different cohorts. Specifically, the component correlating with SARS-CoV-2 incidence in the Netherlands, which is characterized by absence of smell and taste, was detected in both countries. Additionally, we observed in both countries a component dominated by nosebleed and one component with a symptom profile specific of a gastro-intestinal infection.

**Figure 4:**
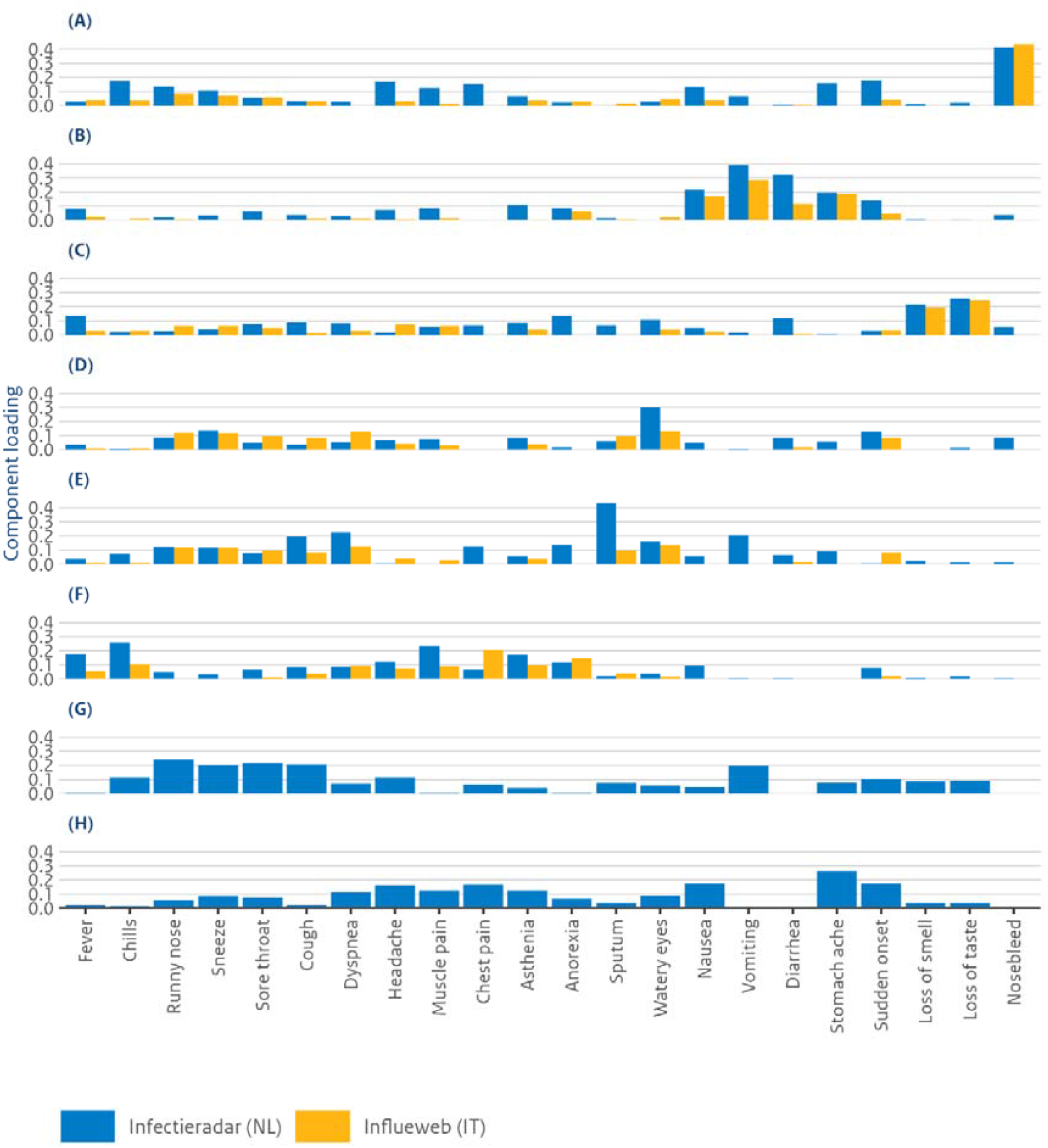
Comparison of the syndromic spectrum of the Dutch and Italian components. NMF was applied separately to the Dutch Infectieradar and Italian Influweb symptom matrix to extract independent syndromic spectra. The component time series of the Dutch components correlated with the following pathogens from submitted nose- and throat swabs of the Dutch Infectieradar participants: (A) undefined, (B) undefined, (C) SARS-CoV-2, (D) undefined, (E) influenza virus, RSV and seasonal coronaviruses, (F) undefined, (G) rhinovirus, and (H) undefined. Note: only six components were detected in the Italian Influweb data.

## DISCUSSION

In this study, we applied NMF to weekly symptom reports from participatory surveillance data to identify latent components that reflect the circulation of specific respiratory pathogens. By aggregating population-level symptom matrices from the Dutch participatory surveillance platform Infectieradar from November 2020 to May 2025 into additive components, NMF revealed distinct syndromic time series, several of which strongly correlated with laboratory-confirmed pathogen incidences. Notably, one component closely tracked SARS-CoV-2 cases, characterized by loss of smell and taste, fever, and cough. Another component was highly associated with rhinovirus incidence, dominated by runny nose and respiratory symptoms, while a third correlated with influenza virus, seasonal coronaviruses, and RSV. Cross-country validation showed consistency in key components between Dutch and Italian cohorts, particularly those associated with SARS-CoV-2 and gastrointestinal infections. These findings demonstrate that unsupervised matrix factorization can extract pathogen-specific signals from weekly symptom data.

A central assumption underlying this approach is that collected weekly symptom reports contain hidden symptom groups that arise from the population-level expression of different respiratory infections. In other words, we hypothesized that each pathogen leaves a characteristic “signature” in the co-occurrence and temporal incidence of symptoms, and that these latent signatures can be recovered through the NMF decomposition. This assumption is supported by clinical evidence that pathogens slightly differ in their typical symptom patterns (14,15). Our results show that NMF can detect these differences at the population level and that the resulting components correlate with laboratory-confirmed pathogen incidence, providing empirical support to our hypothesis. At the same time, our findings leave space for further improvements, given the substantial symptom overlap across respiratory infections (14,16). Nonetheless, the ability of NMF to retrieve meaningful components associated with at least two pathogens suggests that these latent structures are sufficiently stable to be detected in routine surveillance data.

Methodologically, NMF offers clear advantages for syndromic surveillance. Its non-negativity constraint yields symptom loadings that are readily interpretable and biologically plausible. Because NMF leverages temporal covariation of symptoms rather than predefined case definitions, it can detect pathogen-associated patterns even in the presence of high symptom overlap or nonspecific presentations. The generalizability of the syndromic spectra of the components across countries further suggests that underlying symptom–pathogen relationships are relatively stable even within different source populations, opening the possibility of using laboratory data from one well-sampled country to inform symptom-based surveillance in countries with limited or no virological testing.

NMF decomposition of the Dutch dataset yielded eight components, two of which correlated with SARS-CoV-2 and rhinovirus incidence. A third component showed the highest correlation with influenza virus, seasonal coronaviruses and RSV incidence. The temporal pattern of this component closely followed the winter peaks of these pathogens, suggesting that it may capture a shared symptom signature common across several co-circulating seasonal viruses. Because these pathogens often circulate simultaneously and produce highly similar clinical presentations, it is challenging to disentangle their individual contributions using symptom data alone. The high correlations observed across multiple pathogens therefore likely reflect a composite “seasonal respiratory virus” signal rather than pathogen-specific dynamics. This component may thus represent a general winter respiratory syndrome driven by the combined circulation of influenza virus, RSV, and seasonal coronaviruses.

The remaining five components of the Dutch dataset did not match any specific pathogen signal to the same extent. Their presence nevertheless highlights that symptom matrices contain rich underlying structure that goes beyond infection trends alone. These additional components may represent patterns linked to non-infectious respiratory conditions, variations in reporting behavior, or co-circulating pathogens not captured in laboratory testing. Furthermore, it is possible that demographic variation (e.g., differential symptom reporting across age groups) or other causes of variation linked to the same pathogen could result in multiple components per pathogen.

### Limitations and Strengths

This analysis benefits from extensive, multi-year participatory symptom data and linked laboratory-confirmed infections, allowing a rare opportunity to evaluate unsupervised decomposition methods against ground truth. Harmonized European surveillance platforms further enabled cross-country data exchange and assessment of component transferability.

However, several limitations must be acknowledged. First, the inclusion of pandemic years introduces atypical symptom distributions and disrupted virus circulation patterns that may bias component learning. Second, while NMF successfully identified pathogen-associated signals, the magnitude of these components should not be interpreted as an estimate of absolute incidence. Component amplitudes reflect the relative strength of symptom patterns rather than exact infection counts, and thus require caution when used to infer epidemiological burden. Third, without virological confirmation of the Influweb cohort, we cannot definitively validate the time trends of the components in Italy. The national surveillance data might be an imperfect proxy to estimate pathogen circulation among the participants of Influweb given the difference in source populations. Lastly, NMF represents only one class of decomposition approaches; alternatives such as independent component analysis (ICA) may capture additional structure or nonlinear relationships and merit future exploration.

## Conclusion

Our approach demonstrates promise for enhancing respiratory pathogen surveillance. When combined with laboratory calibration in at least one country, this framework offers a scalable path to expanding respiratory surveillance capacity across regions with limited resource availability. As participatory and digital surveillance systems continue to expand, these methods could strengthen early-warning capabilities and enhance preparedness for future respiratory epidemics.

## Data Availability

The datasets used and/or analyzed during the current study are available from the corresponding author on reasonable request.

## DECLARATIONS

### Ethics approval and consent to participate

The samples were collected as part of Infectieradar. The protocol for Infectieradar was approved by the Medical Ethics Review Committee Utrecht (reference number: WAG/avd/20/008757; protocol 20-131) was obtained given the nature of data collection. All participants provided informed consent upon registration, which included agreement to the privacy statement and described the processing of personal data and research results, website security measures taken, and how to file a complaint. Participants were eligible to withdraw from the study at any time. Individuals had to be 16 years or over to be able to participate.

For influweb.it the research was conducted in accordance with Italian Data Protection Authority (Garante per la protezione dei dati personali) regulations on privacy, data collection and treatment, reported here https://www.garanteprivacy.it/web/garante-privacy-en. The Italian Data Protection Authority adheres to the norms set by the European Union’s General Data Protection Regulation (GDPR). Influweb’s data collection process was reviewed and approved by the institutional review board of the ISI Foundation which waived the need for ethics approval as the following applies: all use takes place in compliance with the rules contained in the GDPR; informed consent was obtained online from all participants of the platform at enrollment according to regulations, enabling the collection, storage, and treatment of data, and their publication in anonymized, processed, and aggregated forms for scientific purposes; the website has a “Privacy Statement” section in which the users who decide to enroll in the study can find all the information on who is responsible for the data acquisition and processing in each country. Informed consent was obtained online from all participants enabling the collection, storage, and treatment of data, and their publication in anonymized, processed, and aggregated for scientific purposes. The Influweb website (https://influweb.org/) has a “Privacy Policy” section in which the users who decide to enroll in the study can find all the information on who is responsible for the data acquisition and processing. To ensure privacy and data security, all collected data is pseudonymized—participants’ personal identifiers (e.g., email addresses) are stored separately and are inaccessible to researchers. Survey responses are linked only via an anonymized ID, preserving full privacy. Influweb operates on Google Cloud Platform (GCP) infrastructure in Europe, maintaining GDPR compliance, with continuous backups. Additionally, security measures to protect participant data include hashed passwords, twofactor authentication (2FA) and protection of data in transit by Transport Layer Security v1.3 (“TLSv1.3”).

### Consent for publication

Not applicable.

### Authors contributions

All authors contributed to the design of the study. Data from Infectieradar and Influweb were processed and prepared for analyses respectively by GC and MM. GC conducted statistical analyses, drafted the original version of the manuscript, and edited the final version; MM conducted statistical analyses and collaborated on the manuscript draft; NG collaborated on statistical analysis and the manuscript draft; AJVH and DP supervised the statistical analysis and collaborated on the manuscript draft; All authors reviewed, edited, and approved the final manuscript.

## Acknowledgements

We thank the participants of Infectieradar and Influweb for their weekly contributions and submission of samples. Without their contribution this project would not have been a success. Furthermore, we thank our colleagues who help maintain and run the Infectieradar- and Influweb-cohort and handle the samples. Developing and running these studies has been a huge team effort.

## References

1. Zimmerman RK, Balasubramani GK, D’Agostino HEA, Clarke L, Yassin M, Middleton DB, et al. Population-based hospitalization burden estimates for respiratory viruses, 2015–2019. Influenza Other Respir Viruses. 2022 Nov 1;16(6):1133–40. doi:10.1111/irv.13040

2. Friesema IHM, Koppeschaar CE, Donker GA, Dijkstra F, van Noort SP, Smallenburg R, et al. Internet-based monitoring of influenza-like illness in the general population: Experience of five influenza seasons in the Netherlands. Vaccine. 2009 Oct 23;27(45):6353–7. doi:10.1016/j.vaccine.2009.05.042

3. Smith GE, Elliot AJ, Lake I, Edeghere O, Morbey R, Catchpole M, et al. Syndromic surveillance: two decades experience of sustainable systems – its people not just data! Epidemiol Infect. 2019 Feb 22;147:e101. doi:10.1017/S0950268819000074 PubMed PMID: 30869042; PubMed Central PMCID: PMC6518508.

4. Paolotti D, Carnahan A, Colizza V, Eames K, Edmunds J, Gomes G, et al. Web-based participatory surveillance of infectious diseases: the Influenzanet participatory surveillance experience. Clin Microbiol Infect. 2014 Jan 1;20(1):17–21. doi:10.1111/1469-0691.12477

5. van Noort SP, Codeço CT, Koppeschaar CE, van Ranst M, Paolotti D, Gomes MGM. Ten-year performance of Influenzanet: ILI time series, risks, vaccine effects, and care-seeking behaviour. Epidemics. 2015 Dec 1;13:28–36. doi:10.1016/j.epidem.2015.05.001

6. Peppa M, John Edmunds W, Funk S. Disease severity determines health-seeking behaviour amongst individuals with influenza-like illness in an internet-based cohort. BMC Infect Dis. 2017 Mar 31;17(1):238. doi:10.1186/s12879-017-2337-5

7. Kalimeri K, Delfino M, Cattuto C, Perrotta D, Colizza V, Guerrisi C, et al. Unsupervised extraction of epidemic syndromes from participatory influenza surveillance self-reported symptoms. PLoS Comput Biol. 2019 Apr;15(4):e1006173. doi:10.1371/journal.pcbi.1006173 PubMed Central PMCID: PMC6472822.

8. Tara Smit, Gesa Carstens, Wanda Han, Kirsten Bulsink, Jordy de Bakker, Mansoer Elahi, et al. Flexible and scalable participatory syndromic and virological surveillance for respiratory infections: our experiences in The Netherlands. medRxiv. 2024;2024.04.24.24306278. doi:10.1101/2024.04.24.24306278

9. EpiCentro. RespiVirNet - Sistema di Sorveglianza Integrata (epidemiologica e virologica) [Internet]. [cited 2025 Sep 11]. Available from: https://www.epicentro.iss.it/influenza/respivirnet

10. Corman VM, Landt O, Kaiser M, Molenkamp R, Meijer A, Chu DK, et al. Detection of 2019 novel coronavirus (2019-nCoV) by real-time RT-PCR. Eurosurveillance. 2020;25(3):2000045. doi:doi:10.2807/1560-7917.ES.2020.25.3.2000045

11. Lee DD, Seung HS. Learning the parts of objects by non-negative matrix factorization. Nature. 1999;401(6755):788–91.

12. Kullback S, Leibler RA. On Information and Sufficiency. Ann Math Stat. 1951;22(1):79–86.

13. Boutsidis C, Gallopoulos E. SVD based initialization: A head start for nonnegative matrix factorization. Pattern Recognit. 2008;41(4):1350–62.

14. Geismar C, Nguyen V, Fragaszy E, Shrotri M, Navaratnam AMD, Beale S, et al. Symptom profiles of community cases infected by influenza, RSV, rhinovirus, seasonal coronavirus, and SARS-CoV-2 variants of concern. Sci Rep. 2023 Aug 2;13(1):12511. doi:10.1038/s41598-023-38869-1 PubMed Central PMCID: PMC10397315.

15. Osman M, Klopfenstein T, Belfeki N, Gendrin V, Zayet S. A Comparative Systematic Review of COVID-19 and Influenza. Viruses. 2021 Mar;13(3):452. doi:10.3390/v13030452

16. Dietz E, Pritchard E, Pouwels K, Ehsaan M, Blake J, Gaughan C, et al. SARS-CoV-2, influenza A/B and respiratory syncytial virus positivity and association with influenza-like illness and self-reported symptoms, over the 2022/23 winter season in the UK: a longitudinal surveillance cohort. BMC Med. 2024 Mar 26;22(1):143. doi:10.1186/s12916-024-03351-w

