## Supplemental Figure for "Decomposing Participatory Surveillance Symptom Time Series to Track Respiratory Infections: A Cross-Country Evaluation Using Non-Negative Matrix Factorization"

**SUPPLEMENT**

**Table S1. Pearson correlation of the NMF components and the virological surveillance from Italy**

| **Component** | **Influenza virus** | **SARS-CoV-2** | **Rhinovirus** | **seasonal coronaviruses** | **RSV** |
| --- | --- | --- | --- | --- | --- |
| **1** | -0.04  (p = 0.612) | -0.1  (p = 0.277) | -0.24  (p = 0.044) | -0.2  (p = 0.103) | 0.01  (p = 0.956) |
| **2** | 0.36  (p = <0.001) | 0.6  (p = <0.001) | 0.43*†  (p = <0.001) | 0.06  (p = 0.648) | 0.23  (p = 0.052) |
| **3** | 0.26  (p = 0.002) | 0.44  (p = <0.001) | 0.28  (p = 0.02) | 0.05  (p = 0.652) | 0.18  (p = 0.143) |
| **4** | 0.26  (p = 0.002) | 0.17  (p = 0.06) | 0.1  (p = 0.394) | 0.07  (p = 0.54) | 0.23  (p = 0.061) |
| **5** | 0.08  (p = 0.336) | 0.41†  (p = <0.001) | 0.27  (p = 0.025) | 0.03  (p = 0.804) | 0.18  (p = 0.138) |
| **6** | 0.29  (p = 0.001) | 0.04  (p = 0.648) | 0.19  (p = 0.115) | 0.1  (p = 0.396) | -0.01  (p = 0.923) |
| **7** | 0.6†  (p = <0.001) | 0.45  (p = <0.001) | 0.2  (p = 0.1) | 0.05†  (p = 0.7) | 0.7*†  (p = <0.001) |
| **8** | 0.7*  (p = <0.001) | 0.78*  (p = <0.001) | 0.38  (p = 0.001) | 0.14*  (p = 0.263) | 0.57  (p = <0.001) |
| **Note:** ** indicate components with the highest correlation per pathogen* † *indicate components best matching the Infectieradar virological confirmations* | | | | | |

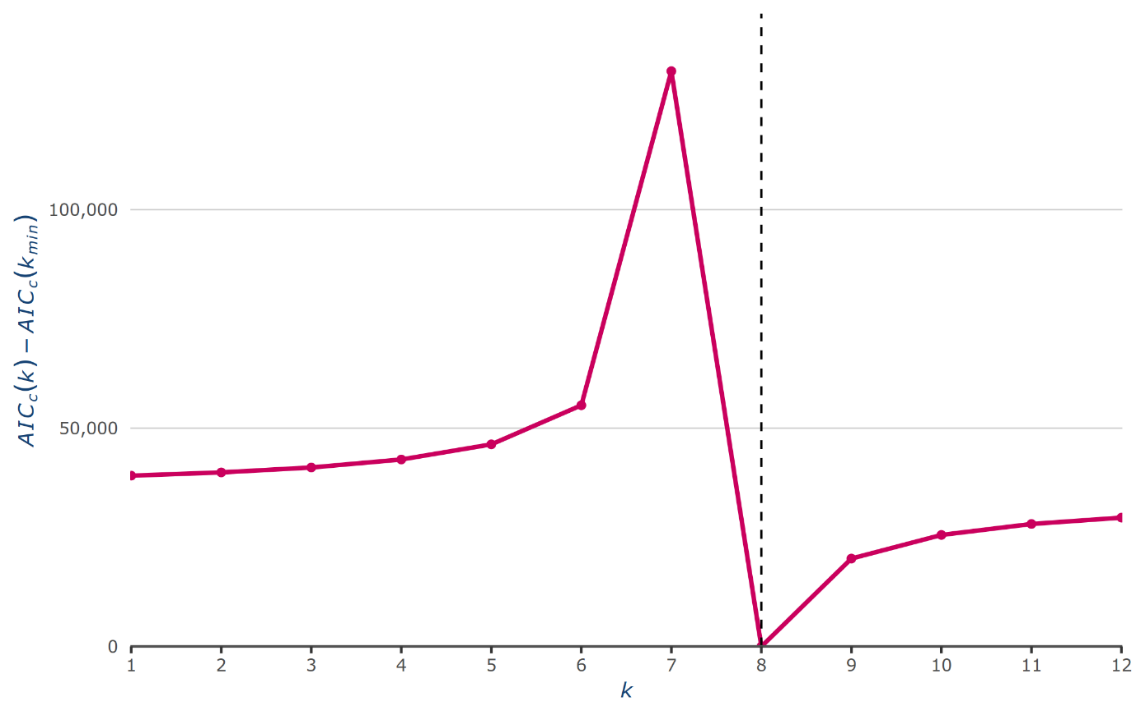

**Figure S1: Selection of the optimal number of components.** To detect the optimal number of components for the Dutch Infectieradar data we calculate for each candidate model the relative likelihood (AIC_c_(k) − AIC_c_(k_min_)) compared against the AIC score of the best model. We depict only models with k < 13 for easier visual inspection. The optimal number of components is visualized with a dashed line.
